# Contracting the Private Health Sector in Thailand’s Universal Health Coverage

**DOI:** 10.1101/2022.06.27.22276979

**Authors:** Aniqa Islam Marshall, Woranan Witthayapipopsakul, Somtanuek Chotchoungchatchai, Waritta Wangbanjongkun, Viroj Tangcharoensathien

## Abstract

**Background:** Private sector plays an import role in health service provision, therefore the engagement of private health facilities is important for ensuring access to health services. In Thailand, two of the three public health insurance schemes, Universal Coverage Scheme and Social Health Insurance, contract with private health facilities to fill gaps of public providers for the provision of health services under Universal Health Coverage. The National Health Security Office (NHSO) and Social Security Office (SSO), which manage the schemes respectively, have designed their own contractual agreements for private facilities.

**Methods:** This qualitative descriptive case study was conducted through document review and in-depth interviews with key informants to better understand the current situation of contracting private health facilities within UHC by comparing the two purchasing agencies on how they contract private primary care facilities, service types, duration of contract, standard and quality requirement and renewal and termination of contracts.

**Results:** Social Security Office (SSO) have adopted capitation-based contract model since 1991. The National Health Security Office (NHSO) applies capitation payments for outpatient services and Diagnostic Related Group systems payment for inpatient services. NHSO classified contractor providers in 3 categories for seamless referral services: main contracting units, primary care units, and referral units. External evaluations and certifications from the Healthcare Accreditation Institute are required for standardization of care. Both purchasing agencies conduct auditing to monitor quality of care while NHSO additionally provides incentives to private facilities for continuous quality improvement.

**Conclusion:** Contracting of private healthcare facilities is needed to fill the gap of public healthcare facilities, especially in urban settings. To ensure access to quality of care, in contracting with private-for-profit providers strong regulatory enforcement and auditing measures with financial incentives in necessary.

## Background

Private sector’s role in health has continued to grow and therefore plays an important role in the provision of health services in many countries.^1,2^ To achieve and advance Universal Health Coverage, engagement of the private sector in ensuring access to health services is crucial.

Thai Universal Health Coverage (UHC) is constructed from a publicly financed health system, consisting of three main public insurance schemes that offer full-service coverage. The three schemes are the Civil Servants Medical Benefits Scheme (CSMBS) for civil servants and their dependents managed by the Comptroller-General’s Department (CGD), Social Health Insurance (SHI) for private sector employees managed by the Social Security Office (SSO) and the Universal Coverage Scheme (UCS) which covers 70.8% of the population in the country managed by the National Health Security Office (NHSO).^3^

Thailand’s health service delivery system is publicly dominant as a result of early and sustained public health system development since 1970s. The public health infrastructure was bolstered through the complete geographical coverage of health centres in all sub-districts, secondary care hospitals in all districts, tertiary care facilities in all provinces and more advanced referral hospitals in all 13 public health regions.^4^ This hierarchical public service delivery system provided a promising platform for successful UHC implementation, with primary care units (defined as district health system contractor networks) acting as gatekeepers for higher level care.^4,5^

However, the district health service delivery system does not exist in major cities, giving room for private clinics and hospitals to fill the primary care gap needed to deliver health services under UHC. Although there are only 382 private hospitals compared to the 2,054 public hospitals in the country, private health facilities contribute 72% of all health facilities.^6,7^ The large majority of these facilities are private clinics which play a significant role in the provision of primary health care under UHC (See table 1). In Bangkok, there are only 69 primary care centres owned by the Bangkok Metropolitan Authority, in contrast to the 6,296 private clinics proving primary care services in the city.

**Table 1.** Numbers of Health Facilities, Hospital Beds and Medical Doctors in the Public and Private Sectors Categorized by Facility Types and Geographic Regions

|  | Public | Private |
| --- | --- | --- |
| Facilities |  |  |
| - Total | 11,959 | 31,286 |
| Hospitals | 2,054 | 382 |
| <90 beds | 814 | 170 |
| 91-500 beds | 221 | 158 |
| 501-1000 beds | 46 | 0 |
| >1000 beds | 11 | 0 |
| No data on number of beds | 0 | 54 |
| Sub-district Health Centres | 9,905 | 0 |
| Clinics | 961 | 30,904 |
| - Bangkok | 261 | 6402 |
| Hospitals | 193 | 106 |
| Bangkok Metropolitan health centres | 68 | 0 |
| Clinics | 0 | 6296 |
| Hospital beds |  |  |
| - Total | 127,275 | 32,566 |
| - Bangkok | 17,051 | 12,594 |
| Medical doctors |  |  |
| - Total | 32,428 | 31,767 |
| - Bangkok | 6,496 | 16,508 |

Source: Ministry of Public Health. Database on Geographic Information System on Health obtained from http://gishealth.moph.go.th/healthmap/gmap.php. Database on Private Hospitals and clinics registered with the Ministry of Public Health's Department of Health Services Support obtained from http://privatehospital.hss.moph.go.th/. Retrieved 11th September 2020

While all public facilities are required to serve as providers for the public insurance schemes, UCS and SHI also engage private providers to serve their urban members where there are gaps in public provider. NHSO and SSO have each designed their own contractual agreements for private facilities that wish to provide services for their beneficiaries. The CGD directly disburses public healthcare facilities for primary care services; and reimburses inpatient care through Diagnostic Related Group system; but does not permit reimbursement of primary care services through private providers.

Although NHSO and SSO contract public and private providers, this study focuses on private contracting—one of the neglected research areas. To better understand the current situation of contracting private health facilities within UHC and identify gaps and challenges, this study compares the two purchasing agencies, namely NHSO and SSO on how they contract private primary care facilities, service types, duration of contract, standard and quality requirement and renewal and termination of contracts.

## Methods

### Scope of Study

This study covered UCS and SHI which have contractual arrangements with private primary healthcare providers.

### Study Design

A qualitative descriptive case study was applied through document review and in-depth interviews with key informants. Data was extracted and analysed into themes based on an adapted conceptual framework for evaluating contracting-out health services.^8^

Ethics approval was obtained from the Institute for the Development of Human Research Protections of Thailand; the certificate number IHRP2020012 on 28 January 2020. Written informed consent was obtained before commencing each interview and key informant information was de-identified to maintain anonymity.

### Context and Setting

Given the private sectors unique role in the city, Greater Bangkok Metropolitan Area was selected as the study setting.^9^ Almost half the private facilities in the country are geographically concentrated in the central region, of which 43% are in Bangkok, where private facilities make up 96% of all health facilities in the city (See table 1). Though most private facilities are clinics or smaller less-than-90-bed hospitals and only 20% of hospital beds in the country are in private facilities, in Bangkok, approximately 42% of all hospital beds are found in private hospitals. Additionally, while the number of medical doctors between the public and private facilities throughout the country is similar, in Bangkok, over 70% of medical doctors work in private facilities.

### Data Collection

Semi-structured in-depth interviews with key informants were conducted to assess the contractual agreements between public purchasing agencies and private providers as well as their experiences and perceptions. Interview guides were separately developed for each of the two stakeholder groups:

First, for purchasing agencies, the major areas of enquiry included: a) profile and characteristics of contracted private facilities; b) contractual requirements, terms, conditions, and regulations; c) effectiveness and challenges in engaging private providers. Stakeholders were recruited through direct request for interviews to the insurance scheme offices. Senior management officers with knowledge on and involvement in implementing contracts with private sector facilities were interviewed.

Second, for key informants from private providers, the major areas of enquiry included: a) general profile and capacity of the health facility; b) requirements, conditions and standards to join public insurance; c) motivations and challenges in contracting with public insurance. Health facilities in the Bangkok Metropolitan Area that have undergone contractual agreements with NHSO and/or SSO were contacted from lists of registered private facilities in the UCS and SHI schemes.^10,11^ Senior management executives and owners of private health facilities with experience in undergoing contractual agreements with NHSO or SSO were interviewed.

Verbal and written consent was obtained from all participants. All interviews were conducted in Thai. Each interview was audio-recorded and detailed field notes were documented. A total of seven key informants were interviewed between Aug 2019-Mar 2020 (See table 2).

**Table 2.** Profiles of Key Informants

| Stakeholder Group | Number and Position | Gender |
| --- | --- | --- |
| Purchasing Agency | 1 Senior Management Officer, NHSO | Female |
|  | 1 Senior Management Officer, Regional NHSO | Male |
|  | 1 Senior Management Officer, SSO | Female |
| Private Provider | 1 Hospital owners | Female |
|  | 1 Hospital management executive | Male |
|  | 2 Clinic owners | 2 Males |

Documents published between 2002-2021, including policies, reports and statistics on private health facilities contracted within the UHC schemes were searched through relevant government agency websites, including the Royal Thai Government Gazette, NHSO, SSO and National Statistical Office (See table 3). Documents were selected based on the title’s relevance to the research topic. Additionally, documents relevant to key informant discussions were requested and obtained directly from key informants. Selected documents were reviewed in full and included if any data applicable to the data analysis themes were identified. Information provided by key informants were verified and triangulated with review of relevant documents.

**Table 3.**
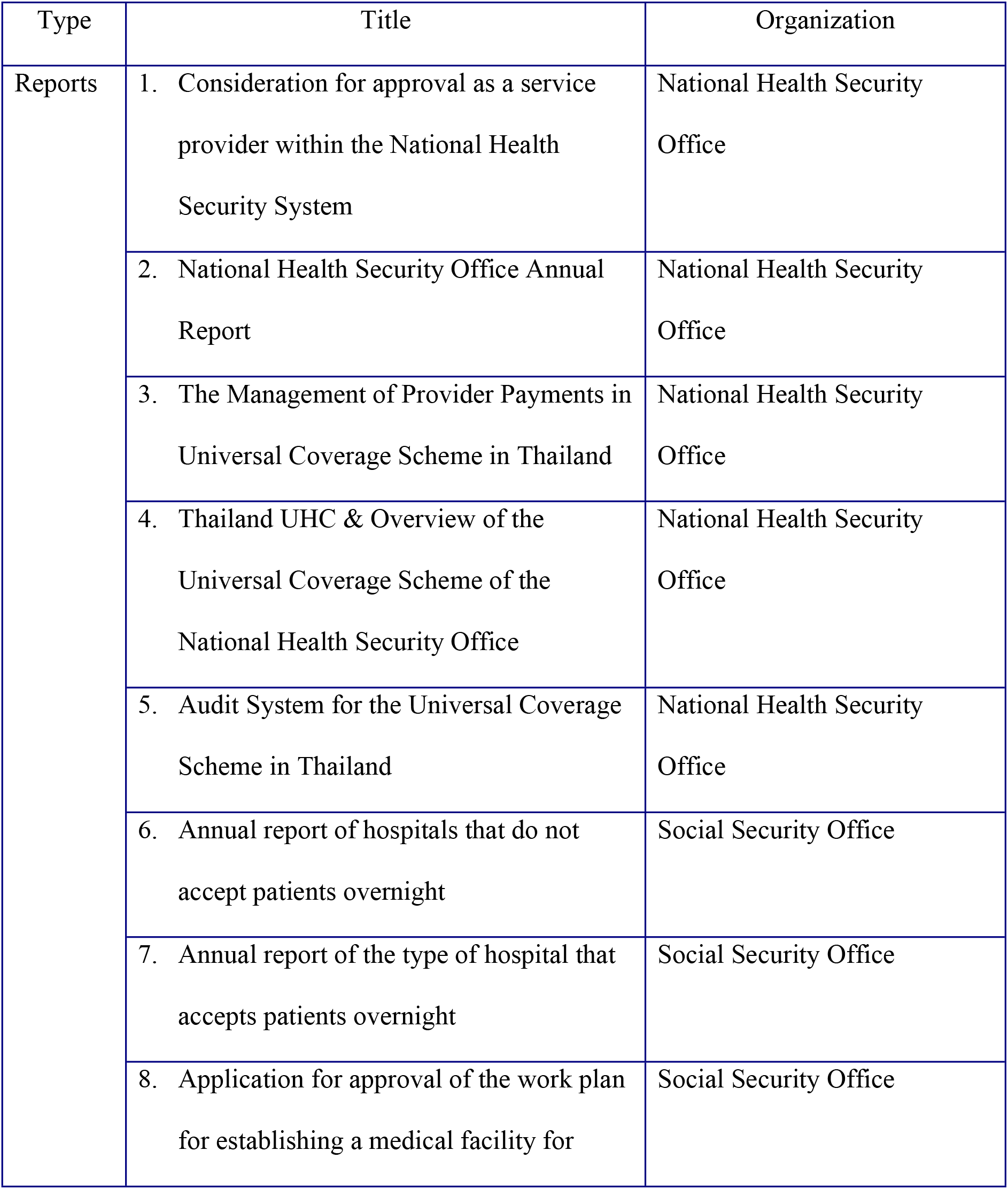
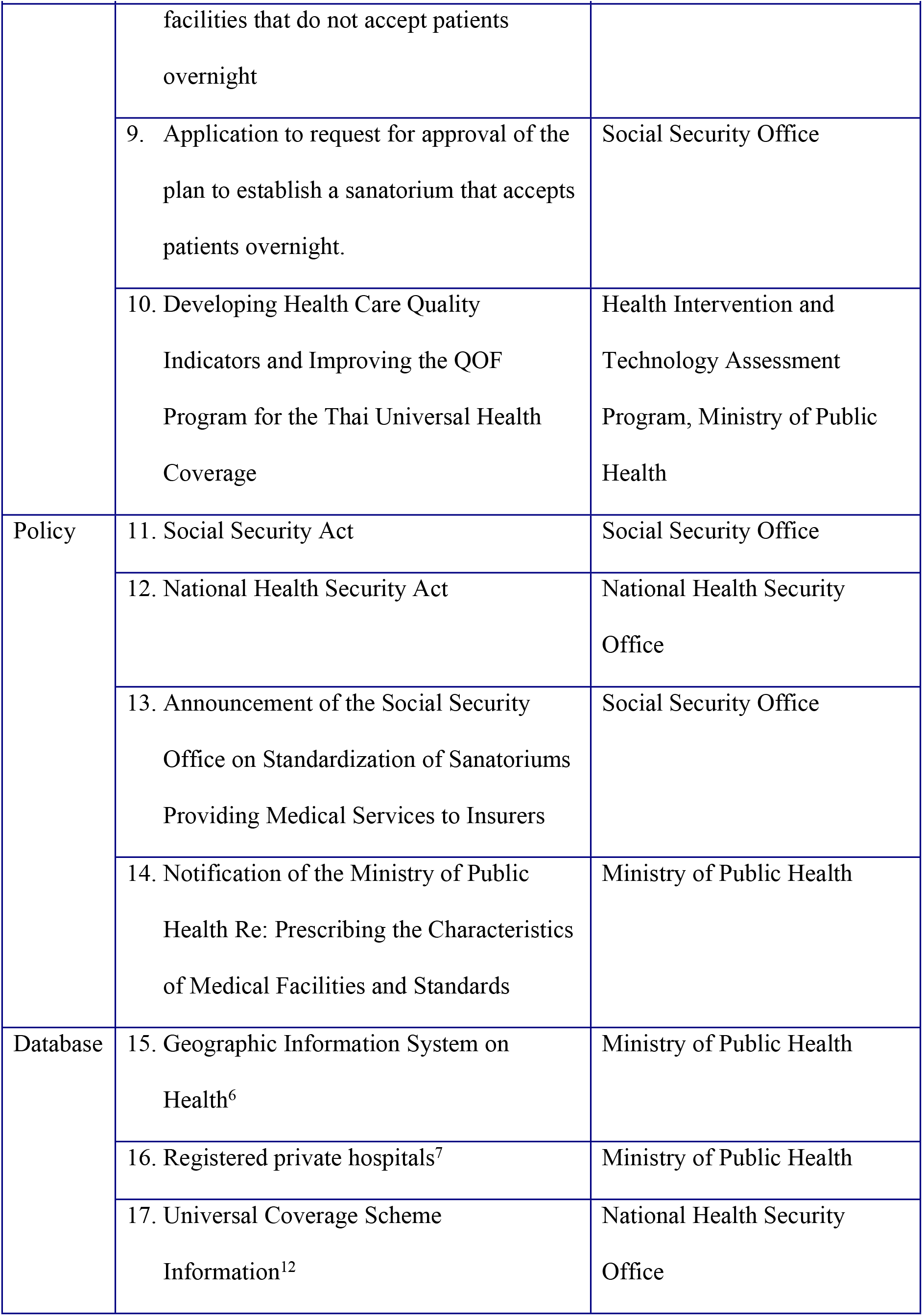
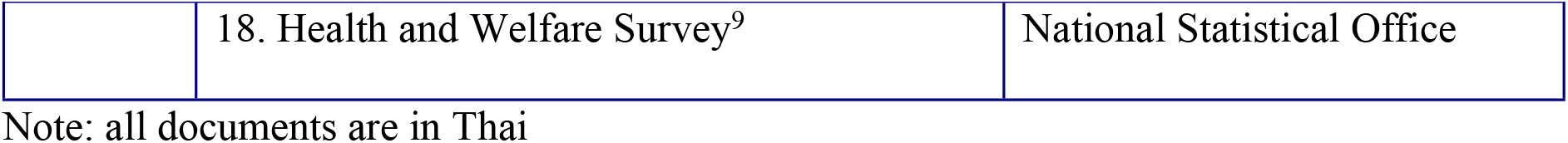
Documents Relevant to Contracting Private Health Facilities in UHC

### Data Analysis

In this study, engagement of private sector is analyzed by the characteristics of the purchaser organizations, the contractor providers, and the contractual relationship, which includes six components: 1) types of services; 2) contract formality; 3) contract duration; 4) provider selection; 5) payment mechanism and 6) performance and quality assurance (See figure 1)

**Figure 1.**
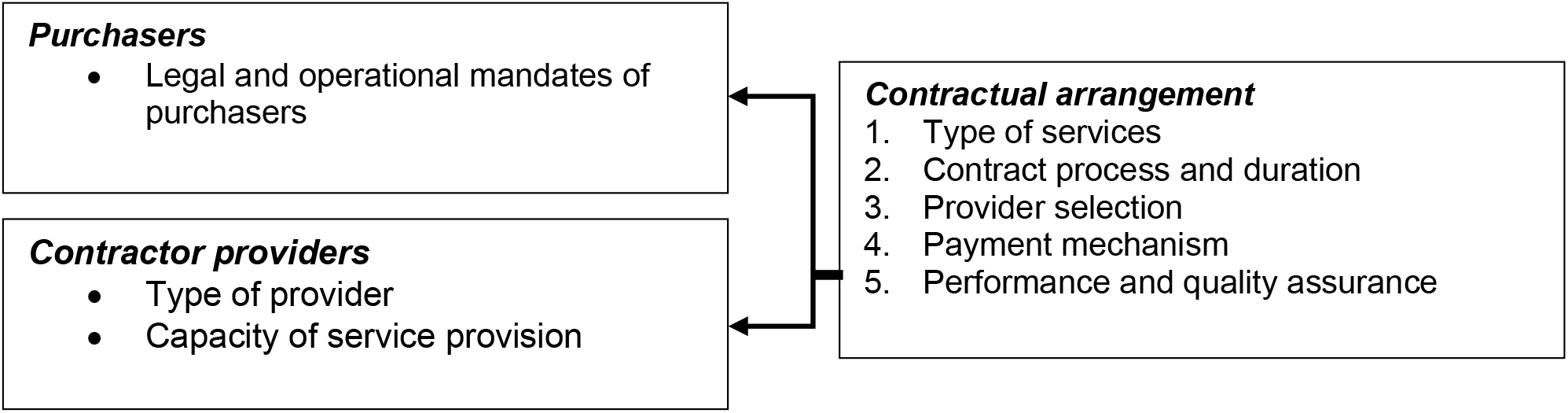
Conceptual Framework for Analysis. Source: adapted from Liu et al; conceptual framework for evaluating contracting-out of health services^8^

Audio recordings from each interview were transcribed and translated. Data from the transcriptions and the field notes were summarized then manually coded and categorized based on relevance to each analysis component. Data obtained from documents were also categorized into each component. Minor quantitative analysis was also conducted by reporting values, ratios and percentages of statistical data obtained from the document review.

## Results

### 1. Nature of Two Purchaser Organizations

In addition to contracting public health facilities, two purchaser agencies, the SSO and the NHSO, are involved in contracting with private health facilities to fill gaps in service provision notably in urban areas.

The SSO, a department in the Ministry of Labour, is a governmental agency, mandated by the 1990 Social Security Act. A tripartite Social Security Committee, consisting of seven government *ex-officio*, seven representatives from employers and seven representatives from employees, is the governing board of Social Security Scheme. The Board is supported by a Medical Committee as advisory role on matters related to Social Health Insurance.^12^

The SSO has several legal mandates. It collects monthly payroll-tax contribution via wire transfer by employers and employees and annual contribution from the government budget allocation, as a tripartite contribution. The Office is required to register all private employers and employees nationwide, maintain database on employee’s salary, as a basis for estimating contributions and provide payments such as for sickness or pension benefits for those retired.^13^

Operating out of provincial social security offices throughout the country and 12 branch offices in Bangkok, SSO contract public and private health facilities on a competitive basis. In addition to medical benefits, the SSO also manages unemployment, child allowance, disability, death compensations and old-age pension as part of a comprehensive social security system.

The NHSO, a public organization established under the 2002 National Health Security Act, is mandated to manage the UCS. The National Health Security Board (NHSB) is the governing body of UCS. Out of the total 30 Board members, five are citizen representatives selected among themselves from the nine civil society organisation constituencies registered with the Ministry of Interior. Responsive governance, reflected by citizen representatives in the governing body, ensures users’ voice and concerns are heard and corrective actions are taken.^14^

The NHSO administers the fully tax-financed National Health Security Fund, through an annual budget negotiation with the Budget Bureau. In addition to the NHSO headquarters, there are 13 regional offices where private sector providers can apply to be registered into the National Health Security System (NHSS) to provide health services to UCS members.

### 2. Contractor Provider Categories

In 2021, the NHSO revised the classification of health facilities that are eligible to join the NHSS into three categories^15^:

- Main Contracting Units which enter into contractual agreement with and are account to NHSO. It has the capacity to provide comprehensive prevention, health promotion and curative services and shall have a referral hospital backup.
- Primary Care Units which cannot enter into contractual agreement with NHSO. It forms into network of primary care units and affiliate with a main contractor unit. It provides services to the catchment population and is paid by the main contracting unit.
- Referral Units who provide treatment for referral cases from the main contracting units and paid by them.

In the past Joint Service Providers were contracted to provide specific services such as dental, rehabilitation and pharmacy services, etc. However, as the interpretation of the revised legal text did not allow for providers to be categorized as “joint service providers”, they are labeled as referral providers in practice.

In 2019, there were a total of 12,337 public and private health facilities registered with NHSO nationwide but less than 5% (580/12,337) were private (of which 301, 281 and 266 were Primary Care Units, Main Contracting Unit and Referrals Units), See Table 4.

**Table 4.**
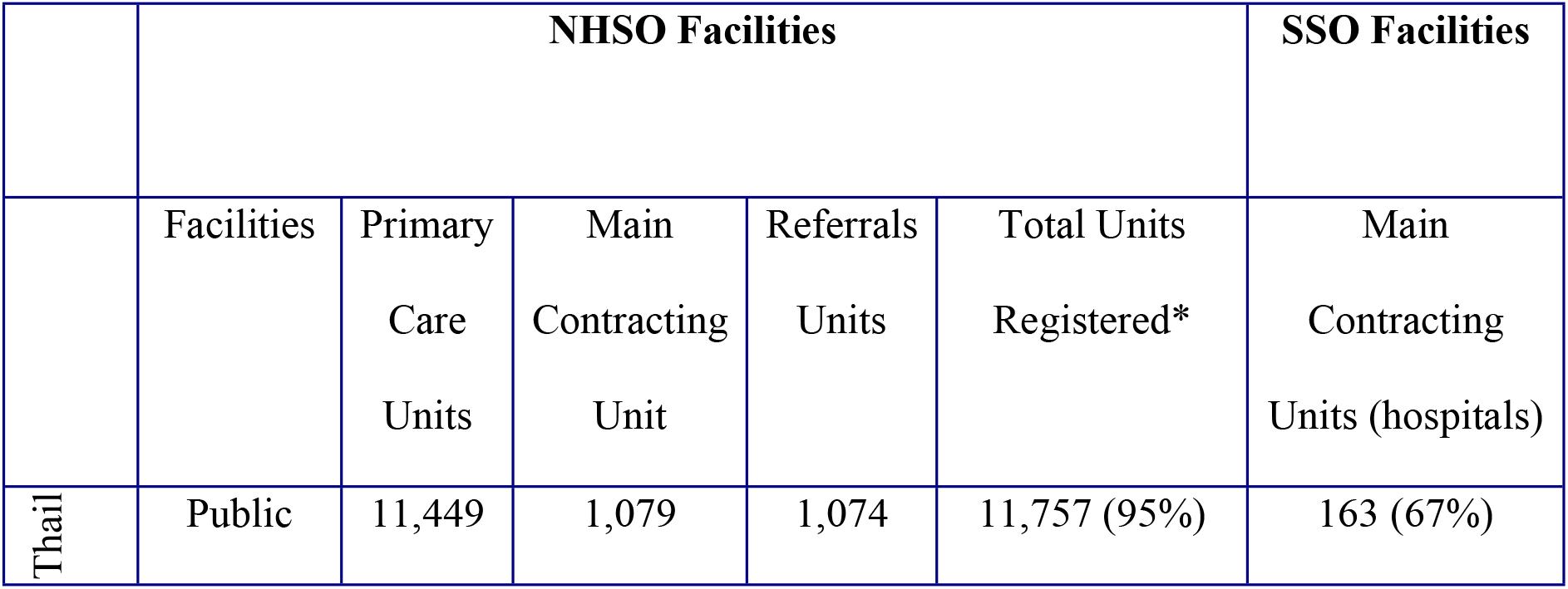
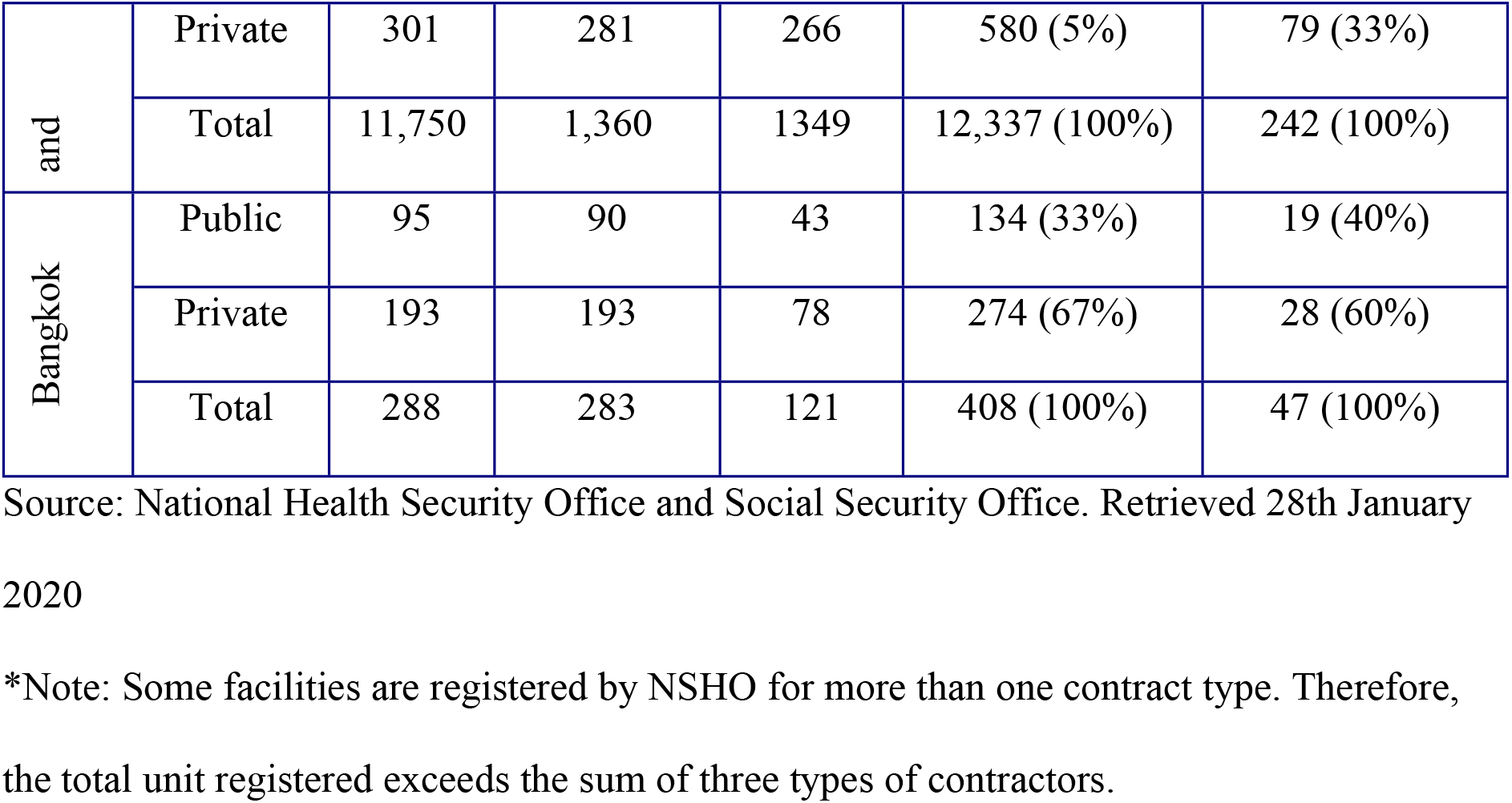
Facilities Contracted within the Universal Health Coverage System in 2019

However, in Bangkok, private facilities made up nearly 70% (274/408) of all facilities providing health services to UCS members. Of these 274 private providers, 193 facilities serve as main contracting units which enter a contractual relationship with NHSO, 193 facilities provide primary care services, and 78 facilities only accept referrals from main contracting units. Note that some facilities register with NHSO as more than one category type. Hence sum of three types of facilities exceeds 274 facilities.

All public and private hospitals with contractual agreements with Social Security Office are called main contracting units; there is no other differentiation. These main contractors are hospitals that satisfy minimum resource and standard requirements set by SSO (such as facilities having at least 100 beds and 12 specialties) and receive direct payments from SSO. Private main contractors are usually equivalent to the size of provincial hospitals with a 150-500 bed capacity.

Applications from private providers to provide health services for SHI members, are assessed at headquarter and provincial branch offices. In 2020, SSO contracted 242 hospitals in the country, with 33% being private. Of all private facilities, 35% are located in Bangkok, where private health facilities constitute almost 60% of all health facilities. Approximately 40% of SHI members in the country and 52% in Bangkok choose to register with a private main contractor hospitals. Although a total of 242 providers for SHI appear small, this number only represents main contractors, and does not include subcontractors and supra-contractors which also provide health services to SHI members.

SSO allows main contractors to enter into agreements with ‘supra-contractors’ and ‘sub-contractors’ which serve in their service network. Supra-contractors are referral hospitals or specialized hospitals that agree to provide services to patients referred from the main contractors when patient needs are beyond their capacity; these are usually regional hospitals, teaching hospitals, or specialized hospitals in the same or neighbouring provinces. Sub-contractors are facilities that support main contractors in providing primary care services, provided service standard are maintained, which can be hospitals or clinics located in the same geographical areas. SSO does not directly contract sub- and supra-contractors, and only the main contractor hospitals are legally accountable to the SSO.

### 3. Contractual Arrangement

#### 3.1 Types of Services

UCS members are entitled to a comprehensive benefits package including disease prevention, diagnosis, medical treatment, health promotion, rehabilitation and dental services, as well as Thai traditional and alternative medicine. In addition to basic inpatient and outpatient services, the NHSO’s targeted and special services include thrombolytic therapy for ischemic stroke patients, cataract lens replacement surgery, transplantations (heart, liver, stem-cell), knee arthroplasty, thalassemia, HIV/AIDS including Anti-retroviral treatment, renal replacement therapy, diabetes and hypertension, chronic psychiatric care and long-term care for elderly.^16^ There are no copayments for UCS members as NHSO fully funds all services.

Similarly, the SSO covers all expenses for medical examination and diagnosis, medical treatment, admission to and treatment in medical establishments, medicine and medical supplies, and ambulatory services. Coverage includes organ transplants, HIV/AIDS, haemodialysis, dental treatment, transplantation (bone marrow, kidney and cornea), and renal failure treatment. SSO contracts with the main contractor hospitals to provide inclusive outpatient and inpatient services using an annual capitation fee plus some additional payments for outpatient and inpatients (see detail in sub-section 3.4). Therefore, contracted hospitals must meet all required capacities.

#### 3.2 Contract Formality and Duration

NHSO renews annual contractual agreements with private main contracting units, on a condition that they maintain NHSO standards and pass annual inspections. Call for new health facility applications are announced annually, with applications assessed at NHSO’s regional offices. NHSO can undergo contractual agreements with all three categories of service providers (as described in section 2), with direct payment mechanisms established for the contracted facilities. A health provider may apply to register as one or more facility type. In practice NHSO primarily contracts with Main Contracting Units; while individual primary care units have direct separate agreements and payment mechanisms with Main Contracting Units outside of NHSOs scope. Contractual documents with private providers are legally binding with financial penalties for non-adherence, while agreements with public providers do not have provisions on penalties.

Contractual agreements between the SSO and private main contracting units are legally binding contracts which are renewed annually with new rates, terms and conditions if applicable. Unlike agreements with public main contracting units where contracts are automatically renewed, private providers must undergo annual re-application, review and re-registration process. Calls for new applications and renewals of the incumbent contractors are announced between April and May through the SSO’s website and invitation letters to previously contracted providers. Interested providers submit their applications for review to SSO headquarter or its provincial branch offices. Private providers that are selected must sign contracts by October to provide services from January to December of the upcoming year. For supra-contractors and/or sub-contractors, SSO is not involved in the agreement process; it is left to internal management of the main contracting units. However, main contractors are required to notify SSO upon application with details about their sub- and supra-contractors’ facility licenses, service scope, and their agreement and consent documents.

New interventions or medicines are introduced into the benefit package and inflation from labour and medical supplies cost can affect the cost of service provision and capitation rate. Both NHSO and SSO apply annual contracts with new rate of capitation and other renewal conditions.

#### 3.3 Selecting Providers for Contracting

A minimum eligibility criterion for a main contracting unit has been established by the NHSO. This includes a) being a health service provider that is already operational, b) must have received a unique health service provider identification number generated by the Bureau of Policy and Strategy, Ministry of Public Health, c) if the health service provider is a primary care unit, it must have an agreement with a host service provider that serves as its main contracting unit, d) if the health service provider is a main contracting unit that is not also registering as a referral unit, it must have a referral unit already registered within the National Health Security System. All NHSO regulations aim for a seamless referral of patients who require secondary or tertiary hospital services.

Additionally, main contracting units must have a doctor to population ratio of 1:10,000, referral units must have a minimum of 30 beds and have at least four main specialties, namely obstetric and gynaecology, general surgery, internal medicine and paediatric, while private sector providers must be licensed and re-licensed annually by the Bureau of Sanatorium and Art of Healing, Ministry of Public Health.

Prior to entry into contractual agreement, the NHSO’s service inspection unit assesses the applicants registering as main contracting units and/or referral units based on a nationally set standards and guidelines developed jointly by NHSO, Ministry of Public Health and relevant stakeholders. Criteria of pre-entry assessment differs by types of contracting units. NHSO also re-assess the contractors annually to ensure these standards are maintained. The one-day assessments are conducted by trained public sector medical doctors and nurses for both new registrations and annual re-assessment inspections. In practice, the main contracting units are inspected and assessed by staff from their affiliated referral hospitals. Facilities that do not pass either the initial inspection or annual inspection are requested to make either general or specific urgent improvements within 7 days otherwise they are removed from the NHSS.

The private main contractors for Social Health Insurance must meet the following requirements: a) having at least 100-bed capacity; b) ability to arrange referrals to ensure patients are treated to the utmost medical care they need; and c) having at least 12 medical specialties [Internal medicines, General Surgery, Ob-Gyn, Paediatrics, Orthopaedics, Preventive Medicines, ophthalmology, anaesthesiology, Ear Nose and Throat, Radiology, Rehabilitation, Emergency medicine or neuro-surgery]. The facilities that do not meet all criteria may be contracted on an exceptional circumstance as advised by the SSO’s medical committee; for example, if it is the only health facility in the underserved areas in particular public contractor. SSO also sets standards for specific areas such as general facility standards, emergency services, outpatient / inpatient / intensive care, surgery, pharmacy, and medical records. For the registration of new applicants, main contractors must also be an accredited hospital either through the Joint Commission International (JCI) or Thailand Healthcare Accreditation Institute (HAI). These requirements and regulations are not enforced on sub-contractors and supra-contractors which are separately contracted by main contracting facilities.

#### 3.4 Provider Payment Mechanisms

Both NHSO and SSO make direct payments to facilities that have undergone direct contractual agreements and implements a blended payment model.

From the NHSO, Main contracting units providing outpatient services to UCS members are paid prospectively one month in advance as well as general health promotion and disease prevention services. The capitation payment is calculated for each facility by multiplying an age-adjusted differential capitation rate with the number of members registered to the main contracting unit; the more number of elderly and children under five years old receives slightly higher outpatient capitation rate. The total budget for outpatient is calculated for each contractor unit based on the population in the catchment area registered with each contractor. Age adjustment is applied to 80% of capitation rate while the remaining 20% is flat rate equally for any main contracting unit.

In-patient services for UCS members are paid through retrospective claim and reimbursement by applying Diagnosis Related Groups (DRGs) System under a global budget.

Reimbursement payments are also used for special health promotion and disease prevention services in cash and in kind for specific services such as vaccines, anti-retroviral drugs and peritoneal dialysis solutions in the form of medicines and medical supplies. Providers are responsible for filing their reimbursement claim forms within 30 days after the inpatient was discharge from hospital, through the NHSO e-Claim program managed by the NHSO Bureau of Information Technology. The claim is then verified by the Bureau of Fund Allocation. Upon satisfactory verification, providers are compensated by the Bureau of Fund Accounting through electronic bank transfer directly to the health facility.

In addition to capitation for outpatient and DRG reimbursement for inpatient, a fee schedule under a global budget is applied for high-cost services that may pose a financial risk to health facilities, such as knee prosthesis and high-cost medication such as recombinant tissue plasminogen activator for the treatment of acute ischemic stroke.

NHSO also applies a fixed fee per patient for some non-communicable diseases screening services, and a point system with global budget for acute diseases, emergency services, Thai traditional medicines and rehabilitation services. Activity-based contracts for area-based and community-based health promotion and disease prevention services to meet the needs of local health problems are managed by regional NHSO offices and paid as project-based “block-grants”. The grant is allocated to sub-district health centres. All of these blended payment to providers aims to improve access to high cost but effective interventions and minimize the weakness of potential “under provision of services” of the main mode of capitation payment.

The Social Security Office applies a mix of payment methods, which include capitation, point-system, DRG, and fee-schedule. Outpatient and inpatient services are paid monthly based on an annual capitation rate of 1,500 THB per SHI member registered to the facility. For inpatient care with an adjusted relative weight over 2 under the DRG systems, services for these patients are paid by applying DRG with global budget. For inpatient with relative weight below 2, it is covered by the 1,500 THB capitation rate.

To incentivize providers to provide adequate treatment for patients with chronic diseases, an additional 453 THB per registered SHI member is allocated with 50% paid monthly, and the remaining 50% paid at the end of annual contract, subject to quality performance earned by a point system. Certain equipment and preventive services can be reimbursed using a fee-schedule. SSO pays an additional 80% of the amount of expense over 1,000,000 THB for high cost service of more than a million Bath. Supra- and sub-contractors are paid directly from main contractor units, not SSO, with rates and terms mutually agreed upon between the main contractor and the supra- and/or the sub-contractors.

#### 3.5 Performance Requirements and Quality of Care

The NHSO has introduced various mechanisms to monitor health facility performance and ensured service quality. The Health Service Standard and Quality Control Board (HSQCB), one of the two governing bodies of the NHSO, sets and monitors the standards and quality of health providers. The Quality Control subcommittee of the HSQCB sets both national and regional standards to ensure applicability to different regional contexts, including criteria for new entry and contract renewal. Health service providers that do not meet the assessment criteria are unable to enter to contract, while facilities that fail annual inspections are subject to dismissal or contracts not renewed. Apart from the new entry and renewal requirements, NHSO also conducts unannounced ‘surprise visits’ to health facilities, usually for facilities suspected to be non-compliant to NHSO requirements based on document audits, complaints received from UCS members, and random checks with users.

In 2014, to increase quality of services, the Quality and Outcomes Framework (QOF) for primary care providers links payments with their performance. The Thai QOF, which aims to reflect the quality of health promotion and disease prevention, primary care, administrative services, and performance of services, consists of six national health service performance indicators set by the NHSO in collaboration with the Ministry of Public Health and Thai Health Promotion Foundation and not more than five regional specific indicators approved by regional committee based on health and quality challenges faced by the region. See Box 1

Box 1 Quality Outcome Framework Indicators, Fiscal Year 2021^17^

**A. National Core Set of Indicator**

1. Percent of population age 35-74 years are screened for diabetes using fasting blood sugar

2. Percent of population age 35-74 years are screened of hypertension

3. Percent of pregnant women having antenatal care before 12 weeks gestation

4. Cumulative percent of women 30-60 years old having cervical cancer screening in the last five years

5. Percent of outpatients covered by rational use of antibiotics for a) acute diarrhea, and b) acute respiratory infection

6. Hospitalization rate of five ambulatory care sensitive conditions (epilepsy, COPD, asthma, diabetes and hypertension)

**B. Samples of Regional Specific Set of Indicators**

1. Percent of children 9, 18, 30 and 42 months old received developmental screening, and percent children detected with delayed developed received treatment within 30 days

2. Percent of home-bound and bed-ridden elderly received home visits

3. Percent of depressive patients have access to psychiatric treatment

4. Percent primary school children who are obese,

5. Percent primary school children received oral health screening

6. Percent of adults 18-59 years who have normal body mass index.

In 2004, The Healthcare Accreditation Institute of Thailand initiated a new stepwise standardization process to recognize hospitals of varying quality standards.^18^ Step one accreditation was granted for facilities to compliance to preventative measures and risk identification systems in place; step two for facilities meeting key Hospital Accreditation standards of quality assurance and quality improvement; and step 3 was granted to hospitals meeting all Hospital Accreditation standards.

To improve quality of services, in 2007 NHSO provided financial incentives to hospitals based on the achievement of HAI accreditation, by earmarking a budget of 0.76 Baht per capita (35.7 million Baht per annum in total) to boost the quality improvement. NHSO applied a scoring system with a maximum of score of 5 for hospitals with full accreditation, score of 2 for those achieving step 2 and score of 1 for those achieving step 1 from the HAI accreditation. Fully accredited primary health care facilities have been entitled to receive a grant of up to 30 Baht per capita multiplied by catchment population it served.^19^

In contrast to the NHSO, the SSO does not proactively incentivise hospitals for continuous quality improvement. However, the Office relies on external evaluations and certifications from the Healthcare Accreditation Institute or Joint Commission International (JCI) as conditions for private hospitals to enter into agreements with SSO. Additionally, SSO conducts assessments and quality control audits, with main contracting units assessed annually.

## Discussion

Ministry of Public Health healthcare facilities have been significant in achieving UHC and resulting in favourable health outcomes; however, private hospitals have also played an important role, especially in provincial cities.^4^ Private hospitals largely contribute to the health provision in Bangkok, 106 out of total 296 hospitals (35.5%). There are no public PHC facilities in Bangkok; with primary care services being provided by 6,296 private clinics and 68 public health centres operating out of hospital outpatient departments (table 1).

Albeit prior studies show inconclusive evidences on the merits of contracting out health services; contracting private providers are effective and should be advocated in developing countries.^20^ Though contracting out improves access and equity in access to health services, there is little evidence on its impacts on quality and efficiency.^21^ This study does not intend to assess the outcome of contracting but provides a detail analysis and rationale behind contracting and how contractual agreement are practiced in Thailand.

SHI is the pioneer in contracting out health services since its inception. In 1990, public and private healthcare facilities had already provided nationwide coverage, therefore, there had been no need for the SSO to operate its own hospitals to provide services to their members. As a result, a capitation-based contract model was adopted. A single capitation rate is paid to both public and private contracted hospitals to provide outpatient and inpatient services in each year.^22^ This simple model has been applied since 1991 with some modifications such as additional payments for inpatient services with higher case mix severity, as measured by the relative weight of >2 under the Diagnostic Related Group system.

Though a contracting model is also applied by NHSO; it does not follow the same mechanism as SSO. NHSO applies a capitation-based contracting model only for outpatient services; while cost of inpatients services are paid by DRG systems. This is to prevent the downside of a single capitation model where healthcare facilities are reluctant to admit patients in order to save costs as outpatient treatment is less costly for facilities.

NHSO developed a more sophisticated definition of a contractor provider than SSO in order to ensure seamless referral services. For example, Main Contracting Units entering into contractual agreements with NHSO must have a referral hospital backup; and Primary Care Units must be affiliated with a Main Contracting Unit. SSO applies a simpler contractual arrangement with the Main Contracting Hospital only. Arrangement and payment between Main Contracting Hospitals and supra-contractor referral hospital and sub-contractor primary healthcare providers are at the discretion of Main Contracting hospitals, with no involvement with SSO; and no requirements to meet the same quality assessment as compulsory for Main Contracting Units.

NHSO negotiates and receives an annual budget per capita for UCS members from the government. As UCS is solely funded by government budget; NHSO has no responsibility to collect contributions from UCS members. This allows NHSO to direct all efforts to its purchasing function.

NHSO and SSO apply quite similar term and conditions for contracts such as quality pre-requites, inspection and approval prior to entry, annual renewal and duration of contract. The payment rate, terms and conditions are equally applied to public and private contractor providers by the two purchasing organizations. SSO main contracting hospitals pass all the quality requirements as they are public or private tertiary hospitals. In contrast, NHSO has a range of contracted providers such as public primary care facilities, private clinics, main contracting unit which are mostly government district or provincial hospitals; therefore, some do not gain approval at the first inspection.^4^

Evidence from a Cochrane review concludes that payment systems do influence primary care physicians’ behaviour.^23^ It suggests that under fee-for-service payments, primary care doctors provide a higher quantity of primary care services compared to capitation and salary-based doctors. Though the impact of payment systems is not robust enough to be used and applied in every policy context.

As SSO and NHSO both apply capitation for outpatient services; our analysis from the 2019 national Health and Welfare Survey shows a lower age-specific (20-60 years old) annual use rate among SHI members, 0.89 visit per capita for SHI members than 1.24 visits for UCS members. Given the same age group, the lower usage rate is probably a result of more limited access to contracted tertiary care hospitals than UCS members who access care from their registered primary care units; in this case sub-district health centres affiliated with district hospitals.

SSO has never terminated contracts though poor performing facilities voluntarily leave the Scheme. Prior to 2019, NHSO has terminated a few private clinics Main Contracting Units in Bangkok due to fraud. However, in 2020, NHSO uncovered a large scandal by 288 private clinics, dental clinics and hospitals; incurring a total loss of 691 million Baht (US$ 23 million) to NHSO.^24^ Cheating is mostly through the use of “ghost patients”, i.e. using citizen ID number of others who are not users of services to claim screening for diabetes, hypertension and provision of dental services. The NHSO filed cases to the criminal court, terminated contracts, banned the facilities from future contracts, as well as reviewed and strengthened the terms and conditions for contractual agreements and improved auditing systems.^15^ Although NHSO requires verification of citizen ID number to verify status of UCS membership, it is time consuming and difficult to verify if these contracted providers use ID number from ghost patients who are also UCS members.

## Conclusions

To fill the public sector healthcare facility gaps in urban settings, insurance funds inevitably have to enter into contractual agreements with existing private healthcare facilities. The private-for-profit contracted providers require the insurance fund’s strong regulatory, enforcement and auditing measures in parallel with financial incentives to achieve the goal of access to quality care and respond to national health goals such as NCD, prevention and health promotion. Though findings from this study are context specific, it contributes to a better understanding on contractual arrangements between private health facilities and public health financing institutions.

## Data Availability

All data has been provided as part of the submitted article.

